# Risk factors for infection, predictors of severe disease and antibody response to COVID-19 in patients with rheumatic diseases in Portugal – a multicentre, nationwide study

**DOI:** 10.1101/2021.10.01.21264428

**Authors:** Ana Rita Cruz-Machado, Sofia C Barreira, Matilde Bandeira, Marc Veldhoen, Andreia Gomes, Marta Serrano, Catarina Duarte, Maria Rato, Bruno Miguel Fernandes, Salomé Garcia, Filipe Pinheiro, Miguel Bernardes, Nathalie Madeira, Cláudia Miguel, Rita Torres, Ana Bento Silva, Jorge Pestana, Diogo Almeida, Carolina Mazeda, Filipe Cunha Santos, Patrícia Pinto, Marlene Sousa, Hugo Parente, Graça Sequeira, Maria José Santos, João Eurico Fonseca, Vasco C Romão

## Abstract

In order to identify risk factors for SARS-CoV-2 infection as well as for severe/critical COVID-19 in rheumatic and musculoskeletal diseases (RMDs) patients, we conducted a multicentre observational nationwide study of adult patients prospectively-followed in the Rheumatic Diseases Portuguese Register – Reuma.pt – during the first 6 months of the pandemic. We further evaluated the development of IgG antibodies against the receptor-binding domain (RBD) of SARS-CoV-2 in patients with RMDs. We used multivariate logistic regression to compare patients with COVID-19 (COVID-19+) with those who did not develop the disease (COVID-19-) and patients with mild/moderate disease with those exhibiting severe/critical COVID-19. COVID-19+ patients were asked to collect a blood sample for IgG testing ≥ 3 months after infection and results were compared with age-, sex- and sampling date-matched controls. Overall, 179 cases of COVID-19 were registered in Reuma.pt in the period of interest (median age 55 (IQR 20); 76.5% females) in a total of 6404 registered appointments. We found that patients treated with TNF inhibitors had reduced odds of infection (OR=0.16, 95%CI 0.10-0.26, p<0.001), severe disease (OR 0.11, 95%CI 0.01-0.84, p=0.010) and seroconversion rates (OR 0.13, 95%CI 0.02-0.91, p=0.040). Tocilizumab was also associated with a reduced risk of COVID-19 (OR 0.15, 95%CI 0.05-0.41, p<0.001). Older age, major comorbidities (diabetes, hypertension, obesity, cardiovascular disease, chronic pulmonary and kidney disease) and rituximab were associated with an increased risk of infection and worse prognosis, in line with previous reports. Importantly, most patients with inflammatory RMDs (86.2%) were able to develop a robust antibody response after SARS-CoV-2 infection, which was linked with disease severity.

## 1. INTRODUCTION

Infection by the novel severe acute respiratory syndrome coronavirus 2 (SARS-CoV-2) has quickly become a global concern following the initial reports of infected patients in Wuhan, China, by the end of 2019 (1). Risk factors for infection and worse prognosis have been extensively documented for the general population, such as older age, cardiovascular and respiratory disease (2),(3). For the special subgroup of patients with inflammatory rheumatic and musculoskeletal diseases (RMDs), whether treated or not with immunosuppressors, information is scarcer, although observational data is increasingly being published in this regard (4),(5),(6) (7) (8).

As immunosuppression might preclude an adequate antiviral response after exposure to SARS-CoV-2, patients under conventional synthetic (cs), targeted synthetic (ts) or biological (b) disease-modifying anti-rheumatic drugs (DMARDs) have initially been hypothesized to be at increased risk for development of coronavirus disease 19 (COVID-19) and, possibly, severe forms of the disease. However, most observational studies published so far, did not show increased vulnerability of patients with RMDs, pointing to a similar cumulative incidence of COVID-19 to that reported in the general population, with most infected patients exhibiting mild-to-moderate forms of the disease (9),(10),(11). Indeed, it seems that anti-cytokine therapy does not increase the risk for infection or severe disease in patients with inflammatory RMDs (11),(12). These drugs may even exhibit a protective role in the event of critical disease, since this is associated with an hyperinflammatory state, in which high serum concentrations of IL-6, IL-1 and TNF have been reported (13),(14),(15). It remains unclear, though, whether patients with inflammatory RMDs, due to the immune dysregulation posed by the underlying disease and to the treatment with immunosuppressive drugs, are able to generate a proper humoral immune response after infection by SARS-CoV-2.

In this study we aimed to: identify risk factors for infection by SARS-CoV-2; assess the clinical outcomes of COVID-19; find predictors for severe and critical disease (including the need for hospitalization, intensive care unit (ICU) admission and/or the occurrence of death); and evaluate the development of IgG antibodies against SARS-CoV-2 in patients with RMDs.

## 2. METHODS

We performed a multicentre observational nationwide study of adult patients with rheumatic diseases prospectively followed in the Rheumatic Diseases Portuguese Register – Reuma.pt – in the first 6 months of the pandemic in Portugal – from March 2 (first reported case in the country) to September 30. Reuma.pt is a real-life-based nationwide observational registry that captures a large part of patients with inflammatory rheumatic diseases and the vast majority of patients treated with biological therapies. In addition, it also includes a protocol for patients with osteoarthritis and other non-inflammatory conditions (16). Since March 2020, the Portuguese Society of Rheumatology made available a novel module to capture information on clinical manifestations, treatment and outcome of rheumatic patients infected with COVID-19. All centres were invited to participate in the study and instructed to register all patients with a confirmed or suspected infection by SARS-CoV-2, regardless of the rheumatic diagnosis. We collected demographic, clinical and laboratory data in order to describe the epidemiological features and clinical outcomes of rheumatic patients in Portugal infected by SARS-CoV-2.

### 2.1 Study population

Inclusion criteria comprised all adult patients with inflammatory or non-inflammatory RMDs followed at Portuguese rheumatology departments and registered at Reuma.pt, with confirmed or suspected infection by SARS-CoV-2 between March 2 and September 30 of 2020. A confirmed case of SARS-CoV-2 infection was defined as a positive RT-PCR on samples obtained from the respiratory tract or positive seroconversion for SARS-CoV-2, regardless of patient’s symptoms. A suspected case of COVID-19 was defined as presence of fever plus at least one other respiratory symptom (dyspnoea, persistent cough, odynophagia, anosmia and/or dysgeusia), or presence of one of the previous symptoms after a contact with a confirmed case (17). Confirmed and suspected cases of COVID-19 were referred in this manuscript as COVID-19+ patients.

In order to evaluate risk factors for SARS-CoV-2 infection, we also included all adult patients with RMDs not fulfilling the definitions of confirmed or suspected case of COVID-19 (COVID-19-), evaluated within the same time frame, i.e., with at least one appointment registered at Reuma.pt over the period of concern. Moreover, a control group of subjects with confirmed SARS-CoV-2 infection without inflammatory RMDs was used to assess differences in the seroconversion rate in regard to patients with inflammatory RMDs.

Inflammatory RMDs considered were divided into two groups: inflammatory joint diseases (juvenile idiopathic arthritis, rheumatoid arthritis [RA], spondyloarthritis, gout, pseudogout, undifferentiated arthritis, polymyalgia rheumatica, RS3PE, adult onset Still disease) and connective tissue diseases/vasculitis (systemic lupus erythematosus [SLE], systemic sclerosis, Sjögren’s syndrome, myositis, undifferentiated connective tissue disease, overlap syndromes, antiphospholipid syndrome and systemic vasculitis). Non-inflammatory RMDs included were osteoarthritis, osteoporosis, Paget’s bone disease and fibromyalgia.

Laboratory results during COVID-19 were collected as close as possible to the time of SARS-CoV-2 diagnosis or initial hospital admission. For patients with repeated laboratory measurements during the clinical course of their infection, the highest measurements of the results of interest were considered.

Medication is presented individually or in immunosuppressive treatment categories defined as: “None”, including NSAIDs/colchicine exclusively; “Glucocorticoids”, whether or not on concomitant treatment with DMARDs; “csDMARDs”, including methotrexate, sulfasalazine, hydroxychloroquine, leflunomide, mycophenolate, azathioprine in monotherapy; “Tumor necrosis factor (TNF) inhibitors”, “rituximab” and “other b/tsDMARDs” including tocilizumab, ustekinumab, belimumab and janus kinase inhibitors (JAKi), either with or without concomitant csDMARDs.

This study was approved by the Ethics Committee of the Lisbon Academic Medical Centre and all patients signed informed consent.

### 2.2 Determination of risk factors for SARS-CoV-2 infection and characterization of COVID-19+ patients

COVID-19+ patients were compared to those who had registered clinical visits in Reuma.pt in the same time frame, but who did not develop COVID (COVID-19-). Demographics, underlying rheumatic disease and disease activity in the last evaluation before the pandemic for COVID-19-patients and at the time of the SARS-CoV-2 infection for the COVID-19+ patients were assessed, as well as comorbidities of interest and medication.

### 2.3 Determination of predictors for severe and critical COVID-19

Demographic and clinical features of patients with mild/moderate COVID-19 were compared with those with severe/critical disease. Mild COVID-19 was defined as symptomatic disease without evidence of pneumonia; moderate disease was considered when clinical and/or radiographic signs of pneumonia existed but room-air SpO2 was ≥ 90%; severe disease was classified as hypoxemic pneumonia and/or need for hospitalization; and critical disease was reserved for cases requiring admission to intensive care unit or resulting in death (18).

### 2.4 Antibody response to SARS-CoV-2 infection in patients with RMDs

Patients were asked to collect a blood sample for antibody testing against SARS-CoV-2 at least 3 months after the resolution of infection. All samples were processed in a single centre. Considering that SARS-CoV-2 neutralizing antibodies that inhibit viral replication in vitro mainly target the receptor-binding domain (RBD) of the virus spike protein (19),(20),(21), we quantified IgG antibodies recognizing the RBD using ELISA (through the assay developed by Florian Krammer et al, a format that received FDA emergency approval in April 2020 and is described in detail elsewhere)(22)(8). Seroconversion was presumed for any titre ≥1:50. Blood samples retrieved were frozen and stored at the Lisbon Academic Medical Centre biobank (Biobanco-IMM) in order to ensure their quality and viability up to the serology processing. A control group in a 1:2 ratio of age-, sex- and sampling date-matched donors from a national serology survey, without inflammatory RMDs or immunosuppression, with prior confirmed SARS-CoV-2 infection, was used to compare the frequency of positive anti-SARS-CoV2 IgG and respective titration. The control group included patients with a wide range of COVID-19 severity, from asymptomatic patients to severe forms requiring hospitalization. Reuma.pt and Biobanco-IMM are both approved by the Ethics Committee of the Lisbon Academic Medical Centre and by the Portuguese Data Protection Authority (Comissão Nacional de Proteção de Dados).

### Statistical analysis

Categorical variables were reported as percentages, whereas continuous variables were expressed as median (IQR). Continuous variables were compared by the use of the Student’s two-tailed t-test or the non-parametric Mann-Whitney U test as appropriate. Categorical variables were compared using Chi-square or Fisher’s exact test. We estimated the odds ratio and 95% confidence intervals (CI) for independent associations between demographic, disease-related and treatment-related variables and the mentioned outcomes, through multivariate logistic regression analysis. Age, sex and variables associated with the outcome in univariate analysis with a p-value < 0.1 were included in the models with a stepwise backward selection methodology. P-values less than 0.05 were considered significant.

## 3. Results

### 3.1 Characterization of COVID-19+ patients

Overall, 174 confirmed and 5 suspected cases of COVID-19 were recorded in Reuma.pt, out of 6404 patients with registered visits in the period of interest (median age 55 (IQR 20) years; 76.5% females). The majority of patients had inflammatory conditions (N=162;90.5%), mostly inflammatory joint diseases (N=111; 62.0%) and 18.5% (30/162) had moderate or high disease activity at the time of infection (table 1 and supplementary table 1 (S1)). A total of 17 (9.5%) COVID-19+ patients had non-inflammatory conditions, and these were older (p=0.001) and had a higher comorbidity burden (p=0.006) than patients with inflammatory RMDs (table S2). More than two thirds of the patients (N=122; 68.2%) were under DMARDs (cs- or b/tsDMARDs monotherapy or in combination) and 71 (39.7%) were on systemic glucocorticoids. Most patients became infected after direct contact with a positive patient (N=118, 65.9%; table S3) and the most common symptoms (cough, fever, malaise) and laboratory abnormalities (lymphopenia, elevated C-reactive protein) were in line with what was expected (table S3). After confirmed or suspected infection, a minority of patients suspended csDMARDs by their own choice or according to their physician advice (35/100, 35.0%), whereas more than half suspended b- or tsDMARDs (23/40, 57.5%). Most patients under glucocorticoids remained on their usual dosage (60/71, 84.5%).

**Table 1.** Patient characteristics according to COVID-19 status.

|  | COVID-19+<br>(N=179) | COVID-19-<br>(N=6225) | p-value |
| --- | --- | --- | --- |
| Female, N (%) | 137 (76.5) | 4144 (66.6) | **0.005** |
| Age (years), median (IQR) | 55 (20) | 56 (22) | 0.542 |
| Caucasian, N (%) | 150 (83.8) | 4172 (67.0) | **<0.001** |
| Smoking (ever), N (%) | 34 (19.0) | 1271 (20.4) | **0.020** |
| Rheumatic disease by subgroup<sup>1</sup>, N (%) |  |  | **<0.001** |
| Inflammatory joint diseases | 111 (62.0) | 5114 (82.4) |  |
| CTD and vasculitis | 51 (28.5) | 1087 (17.5) |  |
| Non-inflammatory diseases | 17 (9.5) | 9 (0.1) |  |
| Rheumatic disease by diagnosis, N (%) |  |  | **<0.001** |
| Inflammatory joint diseases |  |  |  |
| Rheumatoid arthritis | 48 (26.8) | 2430 (39.0) |  |
| Spondyloarthritis | 32 (17.9) | 1526 (24.5) |  |
| Psoriatic arthritis | 20 (11.2) | 942 (15.1) |  |
| Other<sup>2</sup> | 11 (6.1) | 216 (3.5) |  |
| CTD and vasculitis |  |  |  |
| Systemic lupus erythematosus | 12 (6.7) | 477 (7.7) |  |
| Vasculitis | 8 (4.5) | 139 (2.2) |  |
| UCTD | 8 (4.5) | 46 (0.7) |  |
| Systemic sclerosis | 7 (3.9) | 213 (3.4) |  |
| Sjögren's syndrome | 5 (2.8) | 133 (2.1) |  |
| Other<sup>3</sup> | 11 (6.1) | 79 (1.3) |  |
| Non-inflammatory diseases |  |  |  |
| Osteoarthritis | 9 (5.0) | 4 (0.06) |  |
| Other<sup>4</sup> | 8 (4.5) | 5 (0.08) |  |
| Disease duration (years), median (IQR) | 7.1 (11.7) | 10.6 (12.5) | **<0.001** |
| Disease activity<sup>5</sup>, N (%) |  |  | **<0.001** |
| Remission | 59 (33.0) | 1807 (29.0) |  |
| Low | 65 (36.3) | 1061 (17.0) |  |
| Moderate | 20 (11.2) | 1200 (19.3) |  |
| High | 10 (5.6) | 336 (5.4) |  |
| Not applicable<sup>6</sup> | 17 (9.5) | 9 (0.1) |  |
| Comorbidities, N (%)<sup>7</sup> |  |  |  |
| ≥ 1 comorbidity<sup>8</sup> | 77 (43.0) | 1554 (25.0) | **<0.001** |
| ≥ 2 comorbidities | 35 (19.6) | 423 (6.8) | **<0.001** |
| Ongoing immunosuppressive treatment by class<sup>7</sup>, N (%) |  |  | **<0.001** |
| None | 48 (26.8) | 69 (1.3) | **<0.001** |
| Glucocorticoids | 71 (39.7) | 2130 (34.2) | 1.000 |
| csDMARDs | 82 (45.8) | 1613 (25.9) | **<0.001** |
| TNF inhibitors | 24 (13.4) | 2598 (48.4) | **<0.001** |
| Rituximab | 7 (3.9) | 261 (4.9) | 0.722 |
| Other b/tsDMARDs | 9 (5.0) | 717 (13.4) | **<0.001** |

1 - for the multivariate analysis, inflammatory joint diseases were set as the reference category; 2 - other diagnoses: crystal-induced arthropathies (n=4), polymyalgia rheumatica (n=3), juvenile idiopathic arthritis (n=2), adult-onset Still's disease (n=1), RS3PE (n=1); 3 - other diagnoses: antiphospholipid syndrome (n=3), mixed connective tissue disease (n=3), sarcoidosis (n=2); myositis (n=2), overlap syndromes (n=1); 4 - other diagnosis; fibromyalgia (n=3), osteoporosis (n=3), Paget bone disease (n=2); 5 - disease activity in the last evaluation before the pandemic for COVID-19- patients and at the time of the SARS-CoV-2 infection for the COVID-19+ patients; 6 - in case of non-inflammatory diseases. 7 - specification of comorbidities and individual drug treatments can be found in supplementary table S1. 8 - at least 1 comorbidity among arterial hypertension, obesity, diabetes, cardiovascular disease, asthma, chronic obstructive pulmonary disease, chronic kidney disease. b - biological; cs - conventional synthetic; CTD - connective tissue disease; DMARDs - disease-modifying anti-rheumatic drugs; JAKi - *janus kinase* inhibitors; NS - statistically non-significant; NSAIDs - nonsteroidal anti-inflammatory drugs; TNFi - tumour necrosis factor inhibitors; ts - targeted synthetic; UCTD - undifferentiated connective tissue disease.

### 3.2. Risk factors for SARS-CoV-2 infection

In order to assess risk factors for infection by SARS-CoV-2, we compared the demographic and clinical data of these 179 COVID-19+ patients with 6225 controls (COVID-19-) (table 1 and S1). On univariate analysis, COVID-19+ patients were more frequently female, Caucasian, never smokers and had a higher prevalence of cerebrovascular disease and chronic kidney disease. The prevalence of COVID-19 in patients with connective tissue diseases or vasculitis (4.5%) was more than twice that of those with inflammatory joint diseases (2.1%, p<0.001). Further, COVID-19+ patients had a shorter rheumatic disease duration and lower disease activity. Regarding treatment, patients under csDMARDs (n=82/1695, 4.8%) or b/tsDMARDs (n=40/3616, 1.1%) were significantly less likely to become infected than those not receiving any immunosuppressive treatment (n=48/117, 41%; OR 0.073, 95% CI 0.048-0.112, p<0.001 and OR 0.016, 95% CI 0.010-0.026, p<0.001, respectively), albeit the numbers were small in the latter group. Drug subanalysis (table S1) showed that methotrexate, TNF inhibitors (TNFi) and tocilizumab were more frequently used in COVID-19-patients. On multivariate analysis, moderate/high disease activity and use of TNFi or tocilizumab were independently associated with a lower likelihood of developing COVID-19 (Table 2). On the other hand, being Caucasian or having two or more comorbidities were independent risk factors for SARS-CoV-2 infection.

**Table 2.** Multivariate binary logistic regression model for prediction of COVID-19 infection.

|  | OR | 95% Confidence interval | p-value |
| --- | --- | --- | --- |
| Female | 1.240 | 0.821-1.874 | 0.307 |
| Caucasian | 2.548 | 1.419-4.575 | **0.002** |
| ≥ 2 comorbidities | 2.273 | 1.416-3.649 | **0.001** |
| Disease activity |  |  |  |
| Remission/low disease activity | Ref. | - | - |
| Moderate/high disease activity | 0.419 | 0.269-0.654 | **<0.001** |
| RMD subgroup |  |  |  |
| Inflammatory RMD | Ref. | - | - |
| CTD and vasculitis | 1.051 | 0.663-1.667 | 0.833 |
| Non-inflammatory diseases | 50.412 | 10.928-232.549 | **<0.001** |
| Methotrexate | 0.781 | 0.532-1.145 | 0.205 |
| TNF inhibitors | 0.160 | 0.099-0.260 | **<0.001** |
| Tocilizumab | 0.147 | 0.053-0.408 | **<0.001** |

AUC = 0.714 95%CI 0.670-0.757

### 3.3. Predictors of severe/critical COVID-19

Forty-five (25.1%) COVID-19+ patients developed severe/critical disease, requiring hospitalization, 34 (19%) needed oxygen supply, 12 (6.7%) non-invasive ventilation and 4 (2.2%) mechanical ventilation (Table 3). A total of 10 (5.6%) patients died. In addition to supportive treatment, hospitalized patients were most commonly treated with hydroxychloroquine (25, 55.6%), glucocorticoids (19; 42.2%), azithromycin (13; 28.9%) and nonspecific antivirals (12; 26.7%). Thirty-four patients (19.0%) developed at least one complication, most commonly bacterial superinfection and severe acute respiratory illness (Table 3). Two patients (1 with SLE, 1 with RA) were diagnosed with macrophage activation syndrome and 1 patient with primary Sjögren’s syndrome had a non-fatal thromboembolic event.

**Table 3.** Characterization of COVID-19 in patients with RMDs.

|  | Overall<br>N=179 |
| --- | --- |
| **Type of COVID-19 diagnosis, N (%)** |  |
| PCR confirmed | 170 (95.0) |
| Positive Serology | 5 (2.8) |
| Suspected | 4 (2.2) |
| **COVID-19 severity, N (%)** |  |
| Asymptomatic | 17 (9.5) |
| Mild | 29 (16.2) |
| Moderate | 88 (49.2) |
| Severe | 30 (16.8) |
| Critical | 15 (8.4) |
| **COVID-19 Hospitalization care, N (%)** |  |
| Hospitalization | 45 (25.1) |
| Supplemental Oxygen | 34 (19.0) |
| Non-invasive ventilation | 12 (6.7) |
| Invasive ventilation | 4 (2.2) |
| **COVID-19 treatment, N (%)** |  |
| Glucocorticoids | 19 (9.3) |
| Hydroxychloroquine | 28 (15.4) |
| Azithromycin | 16 (8.0) |
| Other antibiotic | 6 (3.1) |
| Lopinavir/ritonavir | 6 (2.5) |
| Remdesivir | 3 (1.2) |
| Tocilizumab | 1 (0.6) |
| Intravenous immunoglobulin | 1 (0.6) |
| **Death from COVID-19** | 10 (5.6) |
| **Other complications<sup>1</sup>, N (%)** |  |
| Acute respiratory distress syndrome | 13 (6.8) |
| Heart failure | 1 (0.6) |
| Bacterial infection | 16 (9.3) |
| Macrophage activation syndrome | 2 (1.2) |
| Thromboembolic event | 1 (0.6) |
| Acute kidney failure | 8 (3.7) |

1 – Each patient could have more than one complication.

Compared to patients with mild/moderate COVID-19, those with severe/critical course were older, had a higher prevalence of arterial hypertension, diabetes, cardiovascular disease and chronic kidney disease (table 4). A higher percentage of patients with connective tissue diseases/vasculitis (29.4%) and non-inflammatory diseases (47.1%) had severe/critical COVID-19, comparing with inflammatory joint diseases (19.8%, p=0.039). Regarding therapy, the proportion of patients under rituximab was higher in severe/critical COVID-19 patients (11.1% vs 1.5%), while treatment with TNFi was associated with less probability of severe/critical COVID-19 (2.2% vs 17%). No differences were found concerning sex, disease activity status before COVID-19, or median time to symptom resolution and negative RT-PCR. On multivariate analysis, age and treatment with rituximab were the only independent factors strongly associated with severe/critical COVID-19 (table 4).

**Table 4.** Comparison of COVID-19+ patients with severe/critical course or mild/moderate disease.

|  | Severe/critical<br>COVID-19<br>(n=45) | Mild/moderate<br>COVID-19<br>(n=134) | Univariate<br>analysis<br>OR (95%CI);<br>p-value | Multivariate<br>analysis*<br>OR (95%CI); p-value |
| --- | --- | --- | --- | --- |
| **Female, N (%)** | 33 (73.3) | 104 (77.6) | 0.79 (0.37-1.72); 0.558 | 0.76 (0.30-1.95); 0.566 |
| **Age (years), median (IQR)** | 70 (22) | 53 (16) | **1.10 (1.06-**<br>**1.13); <0.001** | **1.09 (1.05-**<br>**1.13); 0.001** |
| **Caucasian, N (%)** | 39 (86.7) | 111 (82.8) | 0.34 (0.07-1.52); 0.156 |  |
| **Smoking (ever), N (%)** | 9 (20.0) | 5 (3.7) | 1.14 (0.48-2.74); 0.762 |  |
| **Rheumatic disease by**<br>**subgroup, N (%)** |  |  |  |  |
| Inflammatory joint<br>diseases | 22 (48.9) | 89 (66.4) | Ref. | Ref. |
| Non-inflammatory | 8 (17.8) | 9 (6.7) | **3.60 (1.25-**<br>**10.39); 0.018** | 1.07 (0.57-6.83); 0.284 |
| CTD/vasculitis | 15 (33.3) | 36 (26.9) | 1.69 (0.79-3.61) | 1.64 (0.65-4.14); 0.298 |
| **Disease duration (years),**<br>**median (IQR)** | 9.7 (14.3) | 6.5 (10.0) | **1.04 (1.00-**<br>**1.08); 0.038** |  |
| **Disease activity before COVID-19, N (%)** |  |  |  |  |
| Remission | 16 (35.6) | 43 (32.1) | Ref. |  |
| Low | 9 (20.0) | 56 (41.8) | **0.43 (9.17-1.07); 0.070** |  |
| Moderate | 5 (11.1) | 15 (11.2) | 0.90 (0.28-2.87); 0.853 |  |
| High | 4 (8.9) | 6 (4.5) | 1.79 (0.45-7.19); 0.411 |  |
| Not applicable<sup>1</sup> | 8 (17.8) | 9 (6.7) | 2.39 (0.79-7.26); 0.125 |  |
| **Comorbidities, N (%)** |  |  |  |  |
| No comorbidities | 16 (35.6) | 86 (64.2) | Ref. | Ref. |
| ≥ 1 comorbidity<sup>2</sup> | 29 (64.4) | 48 (35.8) | **3.25 (1.60-6.57); 0.001** | 1.45 (0.63-3.33); 0.384 |
| **Comorbidities (detailed), N (%)** |  |  |  |  |
| Obesity | 11 (24.4) | 27 (20.1) | 1.28 (0.58-2.85); 0.543 |  |
| Arterial hypertension | 21 (46.7) | 28 (20.9) | **3.55 (1.71-7.35); 0.001** |  |
| Diabetes | 10 (22.2) | 3 (2.2) | **13.03 (3.39-50.05); <0.001** |  |
| Cardiovascular disease | 8 (17.8) | 6 (4.5) | **4.80 (1.56-14.75); 0.006** |  |
| Chronic kidney disease | 4 (8.9) | 2 (1.5) | **6.44 (1.14-36.44); 0.035** |  |
| Cerebrovascular disease | 1 (2.2) | 5 (3.7) | 0.59 (0.07-5.16); 0.630 |  |
| Asthma | 2 (4.4) | 0 (0.0) | NA; 0.062<sup>3</sup> |  |
| Chronic obstructive pulmonary disease | 1 (2.2) | 5 (3.7) | 0.59 (0.07-5.16); 0.630 |  |
| Interstitial lung disease | 1 (2.2) | 1 (0.7) | 3.02 (0.19-49.35); 0.438 |  |
| Hyperuricemia | 1 (2.2) | 2 (1.5) | 1.50 (0.13-16.95); 0.743 |  |
| Malignancy | 4 (8.9) | 4 (3.0) | 3.17 (0.76-13.25); 0.114 |  |
| **Immunosuppressive treatment at COVID-19 diagnosis, N (%)** |  |  |  |  |
| None | 16 (35.6) | 32 (23.9) | 1.76 (0.85-3.64); 0.129 |  |
| Glucocorticoids | 18 (40.0) | 53 (39.6) | 1.02 (0.51-2.03); 0.958 |  |
| csDMARDs | 19 (42.2) | 63 (76.8) | 0.82 (0.42-1.63); 0.577 |  |
| TNFi | 1 (2.2) | 23 (17.2) | **0.11 (0.01-0.84); 0.033** | 0.34 (0.04-2.93); 0.324 |
| Rituximab | 4 (8.9) | 3 (2.2) | 4.26 (0.92-19.82); 0.065 | **9.67 (1.67-56.12); 0.011** |
| Other b/ts DMARDs<sup>4</sup> | 0 (0.0) | 9 (6.7) | NA; 0.114 |  |
| **COVID-19 duration (days), median (IQR)** |  |  |  |  |
| Time to symptom resolution | 25 (19) | 17 (23) | 1.01 (0.99-1.03); 0.436 |  |
| Time to negative RT-PCR | 29 (26) | 31 (18) | 1.01 (0.98-1.03); 0.670 |  |

1 – in case of non-inflammatory diseases. 2- at least 1 comorbidity among arterial hypertension, obesity, diabetes, cardiovascular disease, asthma, chronic obstructive pulmonary disease, chronic kidney disease. 3 – Logistic regression not applicable (NA); p-value state for Fisher's test. 4 – Includes tocilizumab (n=5), ustekinumab (n=1), belimumab (n=1), *janus kinase* inhibitors (n=2). CTD – connective tissue diseases; TNFi – tumor necrosis factor inhibitors.
\* Binary logistic regression model for prediction of severe/critical disease

### 3.4 Antibody response after SARS-CoV-2 infection

Out of the 179 included patients, 79 (44%) performed antibody testing, of which 65 (82%) had inflammatory RMDs and 14 (18%) had non-inflammatory RMDs (figure 1, table S4). Blood samples were collected between days 89 and 331 (median time 237, IQR 125 days) after symptom onset or positive PCR test (if asymptomatic). Seventy (89%) patients had positive IgG antibody titres, with a geometric mean titre of 1/1508 (geometric SD factor (GSD) 4.075; range 1/100 to 1/25600) (figure 1A,1B, table S4). Of these, 46 (65.7%) were on cs- or b/tsDMARD therapy (table S4). All but one (a patient with SLE under hydroxychloroquine who developed fever, cough, myalgias and anosmia following a high-risk contact, but who tested negative on RT-PCR) were COVID-19 confirmed cases. Regarding the 9 patients who did not seroconvert, 6 were RT-PCR-positive confirmed cases, 1 tested negative on RT-PCR and 2 did not perform this test. All of the 3 latter suspected cases had a high clinical suspicion based on symptoms and exposure history. This group of patients who did not seroconvert included 1 patient with mild disease (1 SLE under methotrexate and hydroxychloroquine) and 4 asymptomatic patients, including 3 on immunomodulators (hydroxychloroquine, methotrexate and etanercept).

**Figure 1.**
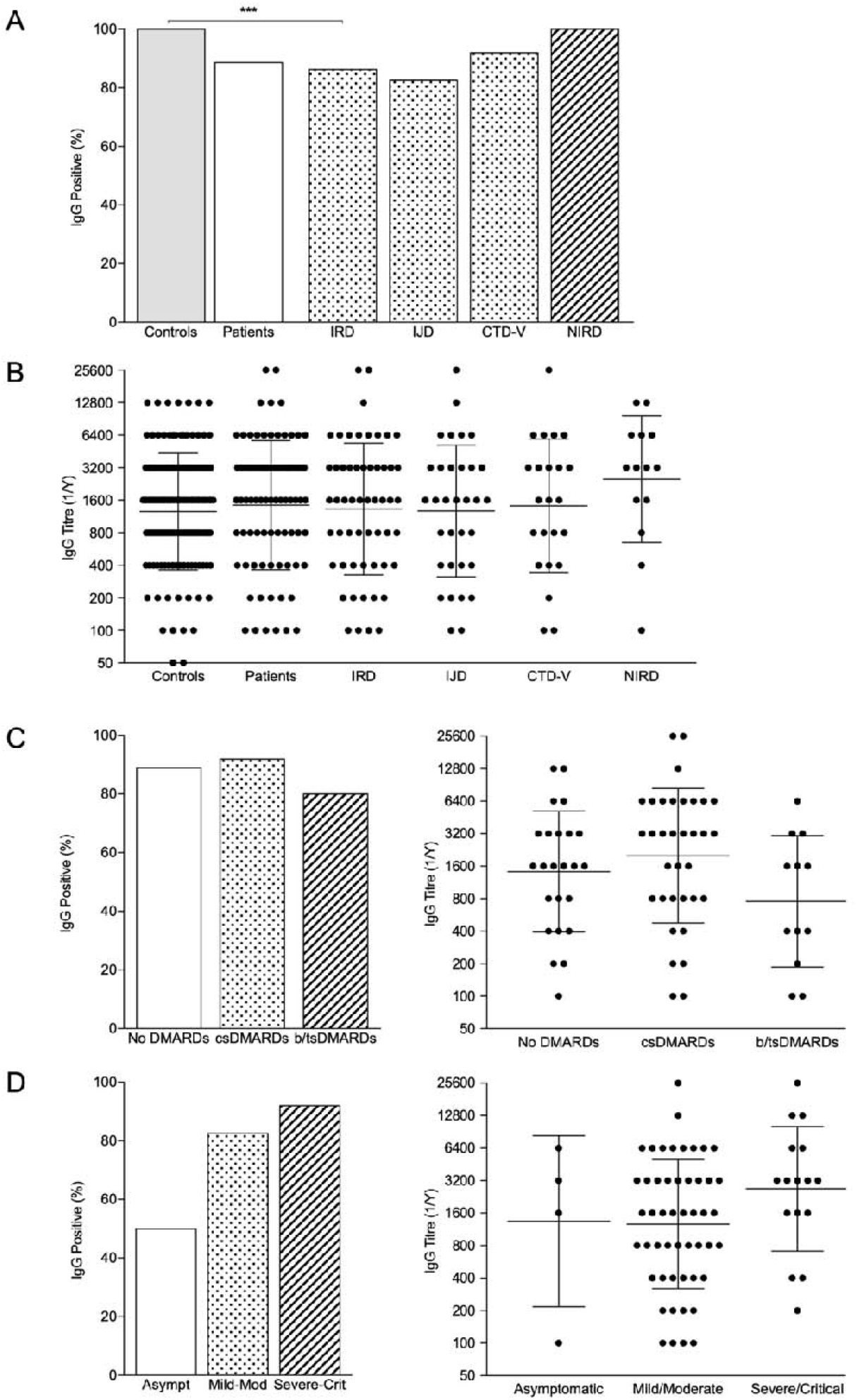
IgG seroprevalence rates and titre distribution across patients and controls. A – proportion of IgG positive patients in controls and patients with individualization per rheumatic disease type. ***p<0.001 B – IgG titres in controls and patients with individualization per rheumatic disease type; bars represent GMT ± GSD. C - proportion of IgG positive patients and IgG titres according to COVID-19 severity; bars represent GMT ± GSD. D – proportion of IgG positive patients and IgG titres across treatment classes; bars represent GMT ± GSD. CTD-v – connective tissue diseases and vasculitis; non-inflammatory rheumatic diseases; DMARDs – disease modifying antirheumatic drugs; IRD – inflammatory rheumatic diseases; IJD – inflammatory joint diseases; CTD-v – connective tissue diseases and vasculitis; non-inflammatory rheumatic diseases

No differences were seen in the seroconversion rate or IgG titres between patients with and without inflammatory RMDs (Figure 1A,1B, table 5, S4). Age, sex and disease activity status at the time of the infection also did not influence seroconversion. Although DMARD therapy as a whole did not influence seropositivity rate, the proportion of patients on TNFi was numerically higher in patients who did not develop IgG antibodies (33.3% vs 8.6%) (Figure 1D, table 5). Of note, all patients treated with corticosteroids (n=30) and rituximab (n=2) developed antibodies. There was no correlation between sample timing and anti-RBD IgG titres (r=0.085, p=0.466). On multivariate analysis, treatment with TNFi and asymptomatic COVID-19 were strongly associated with the absence of serological response (Figure 1D, table 5).

**Table 5.**
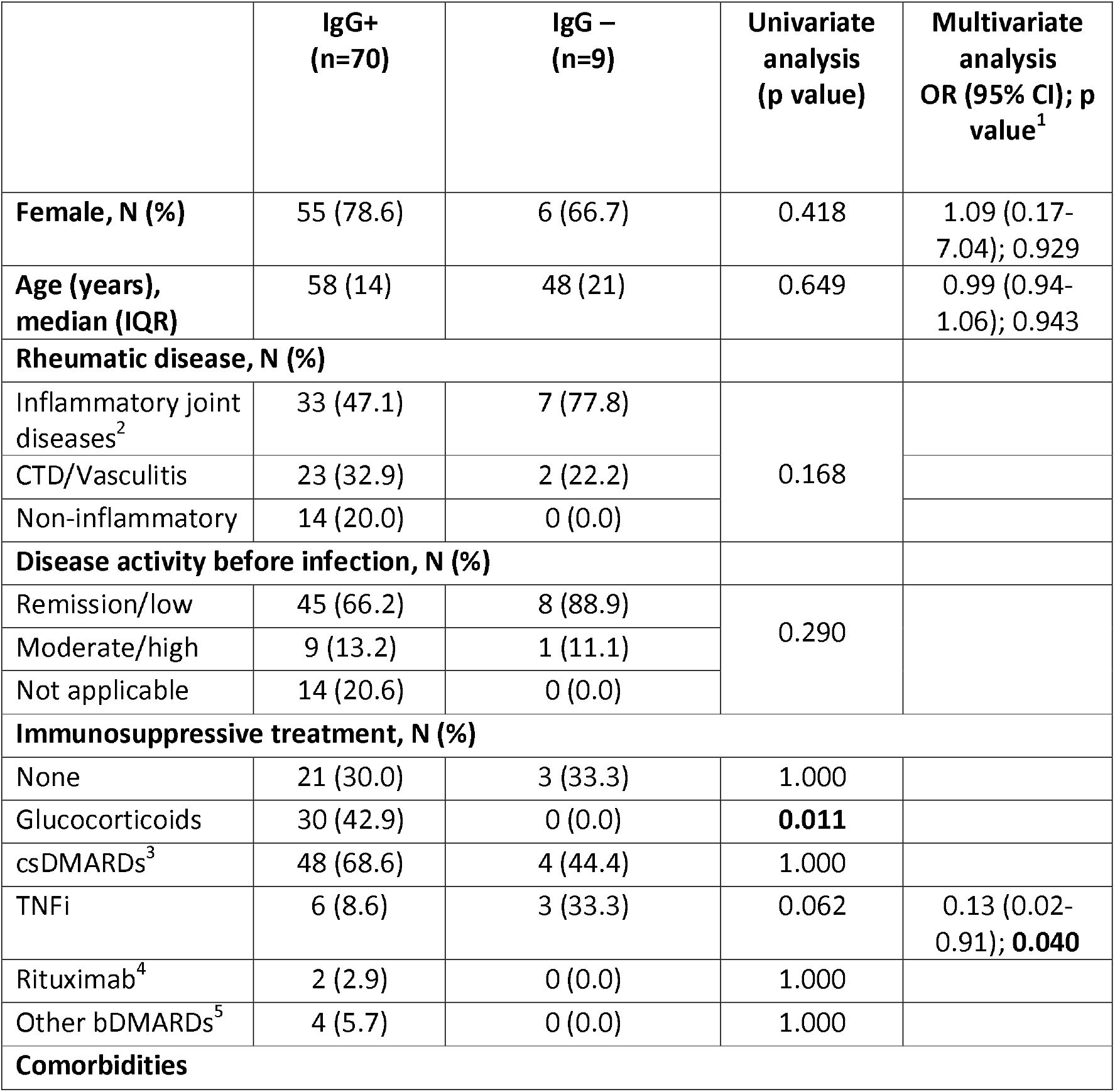
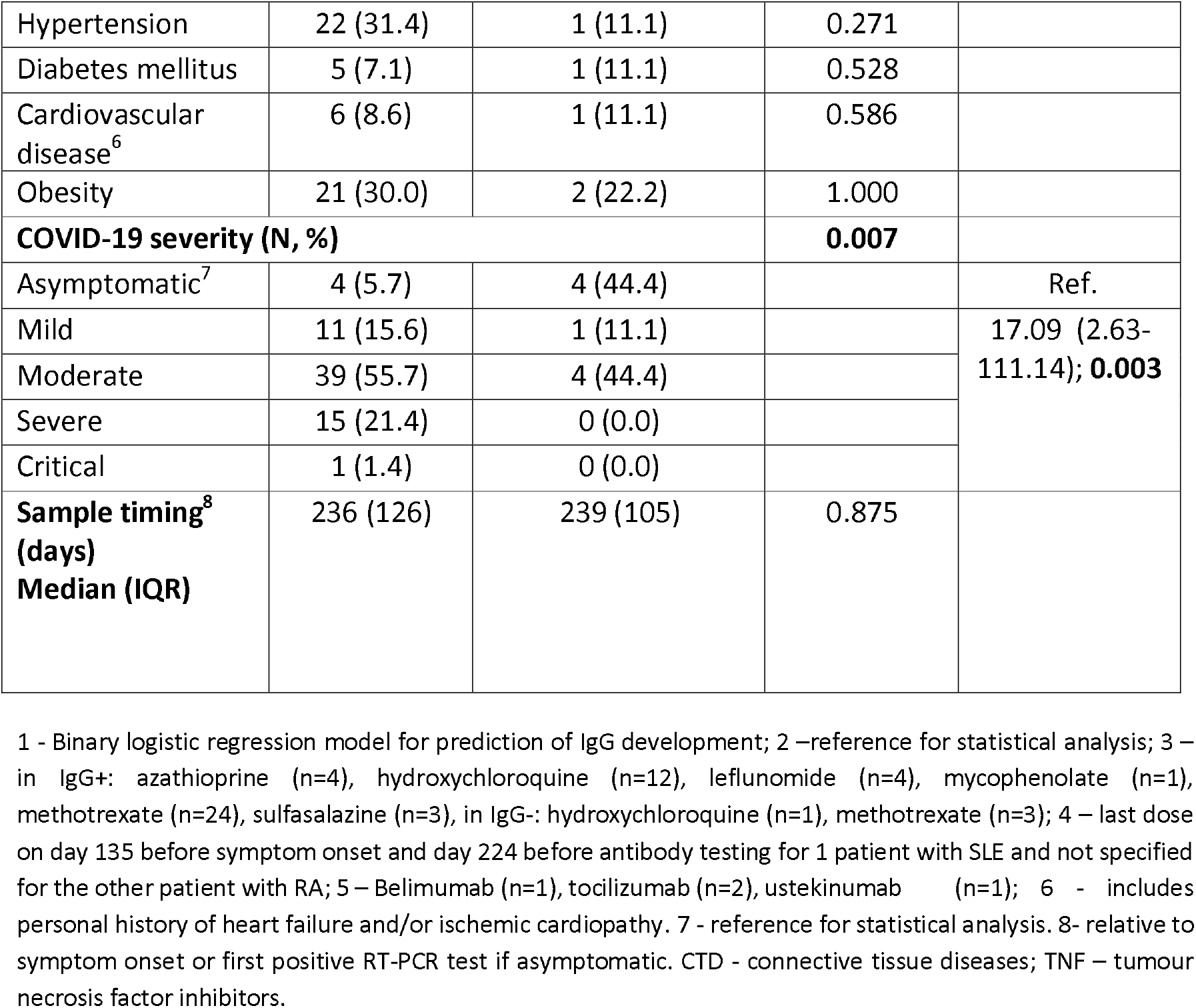
Comparison of COVID-19+ patients based on antibody response.

In comparison with the age- and sex-matched control population (n=130), patients with inflammatory RMDs (n=65) had a lower seroconversion rate (100.0 % vs. 86.2%, p<0.001; Figure 1A, table S5). Nonetheless, antibody titres in subjects with detectable levels did not differ between groups.

## 4. Discussion

During the first months of the pandemic, the European contagion wave hit Portugal with a slight delay, allowing the creation of a dedicated, structured and prospective module in Reuma.pt aiming the assessment of COVID-19 consequences in patients with RMDs. We took advantage of this effort to provide a broad characterization of the SARS-CoV-2 infection in this vulnerable patient population.

We found that Caucasian ancestry and having ≥2 comorbidities were risk factors for infection, while moderate/high RMD disease activity status and treatment with TNFi and tocilizumab were protective. The association of Caucasian ancestry with COVID-19 was unexpected, considering that most studies assessing ancestral and ethnic disparities pointed to a higher prevalence of the disease in Hispanics and Blacks, possibly related to a lower socioeconomic status and a higher comorbidity burden (23),(24). It is possible that other social, cultural, and genetic factors of the Portuguese population contribute to this discrepancy. Regarding disease activity, we may argue that patients with RMDs with more active disease were more aware of the disease itself, as well as of the associated immune dysregulation and drug-related immunosuppression. Thus, they might have complied with measures of social distancing and hygiene in a more scrupulous way. However, it is not possible to exclude a true protective role of some DMARDs, namely TNFi and tocilizumab. Indeed, this is an exciting finding, that is reported in a large population of RMD patients treated with TNFi (n=2622). These targeted therapies are used to treat both inflammatory diseases with more aggressive phenotypes, and severely ill patients with COVID-19, considering the major contribution of TNF and IL-6 to the cytokine-storm syndrome observed in critical patients (13),(14),(25). Similarly to our findings, a smaller single centre retrospective study reported that COVID-19+ patients were less likely treated with IL6-inhibitors (26). Likewise, Simon and colleagues (27) reported a significantly lower SARS-CoV-2 seroprevalence in 534 patients with immune-mediated inflammatory diseases (IMIDs; 295 of whom had RMDs) receiving targeted anti-cytokine therapies compared to healthy individuals (RR 0.32; 95% CI 0.11–0.99). Instead, patients with IMIDs not receiving immunosuppressors exhibited similar seroprevalence to controls (27). We have expanded these results in a considerably larger population (n=3622 RMD patients treated with b/tsDMARDs)

As of September 30, the overall rate of COVID-19-associated hospitalization and mortality in Portugal were, respectively, 8.4% and 2.6% (28). In comparison, the population with RMDs exhibited a strikingly higher burden of the disease — 25.1% were hospitalized and 5.6% died. Although a report bias must be taken into account, this greater risk for worse outcomes may also be explained by a compromised health status and iatrogenesis posed by some baseline therapies. Rituximab is particularly relevant in this regard, as it was the only DMARD independently associated with the risk of severe/critical COVID-19 in our analysis, in line with previous studies (29),(30),(5). In the opposite direction, and in addition to the discussed beneficial effect in reducing COVID-19 risk, TNFi were negatively associated with severe/critical disease, suggesting a potential protective role against the combined outcome of hospitalization and/or death. This observation, though, was not confirmed in the multivariate model. Nevertheless, our results point in the same direction as those advanced by the Global Rheumatology Alliance (GRA) landmark paper including 600 COVID-19+ patients from 40 countries, 46% of whom required hospitalization (4). Therein, older age, comorbidities and prednisone-equivalent ≥10 mg/day were predictors for hospitalization, whereas b/tsDMARD monotherapy showed a protective role, an effect mainly driven by TNFi. Finally, contrary to previous reports (5,31,32), we could not confirm a deleterious prognostic role of glucocorticoids and higher disease activity. This may be attributed to other relevant factors such as disease and treatment heterogeneity between studies, variegated COVID-19 treatment protocols, as well as potential genetic and environmental characteristics that differ across populations.

Concerning antibody response, to the best of our knowledge, ours is the largest collection of patients with RMDs submitted to anti-SARS-CoV-2 antibody testing, excluding seroprevalence studies. The majority of our patients mounted an appropriate IgG response 3 to 11 months post-infection. This is remarkable, considering that most of them were on glucocorticoids and/or DMARDs, including two patients on rituximab. Nonetheless, seroconversion rates were still slightly lower than in controls, albeit there were no differences in antibody titres. These results are reassuring, since they show that having a RMD disease or being on immunomodulators does not seem to greatly influence humoral immune response against SARS-COV-2. Previous reports with limited sample sizes are in agreement with these findings. In fact, 10 out of 13 (76.9%; n=6 on immunomodulators) patients with different RMDs tested positive for anti-SARS-CoV-2 antibodies 7 to 216 days post-infection in a publication by D’Silva et al (33). Of note, 2 out of 3 patients on rituximab had undetectable or variable SARS-CoV-2 antibodies. On the other hand, a seroprevalence study reported that 24 out of 29 (83%) SLE patients with previously confirmed COVID-19 had positive anti-SARS-CoV-2 IgG antibody (34). Most of them were also on immunomodulators (63%), including 3 patients on rituximab. The authors were able to find a positive correlation between Hispanic ethnicity and seropositivity, while other demographic variables, SLE-specific factors and treatment were not associated with SARS-CoV-2 antibody response. In our study, however, we did find that TNFi decreased the odds of seroconversion, while symptomatic disease was associated with a higher likelihood of developing antibodies. A deleterious effect of TNFi on humoral response to vaccines such as hepatitis B virus has been reported in patients with RMDs (35,36). As discussed, TNF is among the major players in the immune response against SARS-CoV-2. Further, a more robust antibody response has been associated with more severe forms of COVID-19 (19). As such, and in line with our results, targeted anti-cytokine therapies, such as TNFi, may, on the one hand, be protective of infection and worse disease outcomes, but, on the other, preclude a proper humoral response (19),(27).

Moreover, although we did not perform serial antibody testing, we were able to show that sample timing did not influence seroconversion or antibody titres, with titres as high as 1/25600 up to 331 days after disease onset. Even though it remains unclear what is the optimal antibody titre to protect against reinfection, and what is the role of T-cell-mediated response in adaptive immunity, our results look optimistic regarding the immune response obtained by patients with RMDs. They suggest that it is not worse than the one observed in the general population (37)(38) (8). Indeed, we demonstrated that anti-SARS-CoV-2 antibodies may persist in patients with RMDs treated with DMARDs for at least up to 11 months. This is close to the 13 months recently advanced by Gallais et al (39), who longitudinally measured specific antibodies in 1,309 healthcare workers.

Our study has some limitations, namely concerning the relatively small sample of COVID-19+ patients included. As such, the aforementioned findings must be interpreted with caution. Genetic or environmental factors specific to the Portuguese population might also preclude external generalization. Also, we did not take into consideration potential different social behaviours of COVID-19+ and COVID-19-patients while assessing risk factors for infection. Finally, ours is not a population-based study, thus we cannot affirm the total number of patients with RMDs affected by COVID-19 in the whole country during the first 6 months of the pandemic.

In conclusion, we conducted a multicentre, nationwide, comprehensive evaluation of COVID-19 in patients with RMDs, aiming to assess risk factors for infection, predictors of severe/critical disease and antibody response. We found that TNFi reduced the risk for infection, the odds for severe forms of the disease, and the likelihood of seroconversion. Tocilizumab also reduced the risk for COVID-19. These findings warrant further confirmation in independent cohorts. On the other hand, older age, general comorbidities and rituximab are associated with increased risk for infection and worse prognosis, in line with previous reports. At last, most patients with inflammatory RMDs seem to be able to develop a proper antibody response after COVID-19, particularly if they had experienced symptomatic disease. Our findings are overall reassuring for patients with RMDs, albeit particular caution must be taken in the more vulnerable patient groups.

## Data Availability

Data referred to in the manuscript are available upon request to the corresponding author.

## Acknowledgements

We would like to acknowledge Merck Sharp & Dohme for funding this entire project. Regarding funding sources of the laboratory, we also would like to acknowledge Sociedade Francisco Manuel dos Santos and Grupo Jerónimo Martins, the European Union Horizon 2020 research and innovation program and Fundação para a Ciência e a Tecnologia. The funders had no role in study design, data collection and analysis, decision to publish, or preparation of the manuscript.

We acknowledge the generous sharing of the expression constructs by Dr Florian Krammer, Icahn School of Medicine at Mount Sinai, New York, USA (Development of SARS-CoV-2 reagents was partially supported by the NIAID Centres of Excellence for Influenza Research and Surveillance (CEIRS) contract HHSN272201400008C), and the protein production by Drs Paula Alves and Rute Castro at Instituto de Biologia Experimental e Tecnológica (iBET) Oeiras, Portugal as part of the Serology4COVID consortium.

We acknowledge all rheumatologists who have participated in data collection through Reuma.pt. We also acknowledge patients for their willingness to cooperate throughout the research in such a troubled time.

