## Supplementary table 1 for "Risk factors for infection, predictors of severe disease and antibody response to COVID-19 in patients with rheumatic diseases in Portugal – a multicentre, nationwide study"

**Supplementary table 1 – Comparison of comorbidities and treatment comparison between COVID-19+ and COVID-19 - patients.**

|  | **COVID-19+ (N=179)** | **COVID-19- (N=6225)** | **p-value** |
| --- | --- | --- | --- |
| **Comorbidities, N (%)** |  |  |  |
| Arterial hypertension | 49 (27.4) | 872 (14.0) | 0.204 |
| Obesity | 38 (21.2) | 660 (10.6) | 0.562 |
| Cardiovascular disease | 14 (7.8) | 207 (3.3) | 0.183 |
| Diabetes | 13 (7.3) | 193 (3.1) | 0.225 |
| Malignancy | 8 (4.5) | 116 (1.9) | 0.539 |
| Chronic obstructive pulmonary disease | 6 (3.4) | 47 (0.8) | 0.060 |
| Cerebrovascular disease | 6 (3.4) | 43 (0.7) | **0.043** |
| Chronic kidney disease | 6 (3.4) | 32 (0.5) | **0.014** |
| Hyperuricemia | 3 (1.7) | 28 (0.4) | 0.226 |
| Interstitial lung disease | 2 (1.1) | 78 (1.3) | 0.443 |
| Asthma | 2 (1.1) | 55 (0.9) | 0.768 |
| **Ongoing treatment, N (%)** |  |  |  |
| NSAIDs | 30 (16.8) | 1077 (17.3) | 0.297 |
| Glucocorticoids | 71 (39.7) | 2130 (34.2) | 1.000 |
| Hydroxychloroquine | 26 (14.5) | 760 (12.2) | 0.913 |
| Methotrexate | 65 (36.3) | 2422 (38.9) | **0.020** |
| Sulphasalazine | 15 (8.4) | 560 (9.0) | 0.454 |
| Leflunomide | 11 (6.1) | 396 (6.4) | 0.662 |
| Azathioprine | 6 (3.4) | 128 (2.1) | 0.45 |
| Mycophenolate mofetil | 1 (0.6) | 96 (1.5) | 0.376 |
| TNFi | 24 (13.4) | 2598 (41.7) | **<0.001** |
| Tocilizumab | 5 (2.8) | 374 (6.0) | **0.024** |
| Rituximab | 7 (3.9) | 261 (4.2) | 0.722 |
| Ustekinumab | 1 (0.6) | 31 (0.5) | 1.000 |
| Belimumab | 1 (0.6) | 29 (9.5) | 1.000 |
| JAKi | 2 (1.1) | 86 (1.4) | 1.000 |
