## Supplementary table 5 for "Risk factors for infection, predictors of severe disease and antibody response to COVID-19 in patients with rheumatic diseases in Portugal – a multicentre, nationwide study"

**Supplementary table 5 – Comparison of patients with inflammatory RMDs and controls regarding seroconversion.**

|  | Inflammatory RMD  (N=65) | Controls  (N=130) | p-value |
| --- | --- | --- | --- |
| Age (years), median (IQR) | 55.0 (17.0) | 55.0 (21.0) | 0.112 |
| Female, N (%) | 48 (73.8) | 76 (58.5) | 0.057 |
| Sampling time*, median (IQR) (days) | 239.0 (143.5) | 181.0 (58.5) | 0.103 |
| Seroconversion, N (%) | 56 (86.2) | 130 (100.0) | **<0.001** |
| IgG  Titers (GM±GSD)  (min.-max) | 1/1329±4.063  (1/100 – 1/25600) | 1/1256±3.464  (1/50- 12800) | 0.191 |

*days after symptom onset or positive PCR test if asymptomatic. GM: geometric mean; GSD: geometric SD factor.
