## Supplementary table 4 for "Risk factors for infection, predictors of severe disease and antibody response to COVID-19 in patients with rheumatic diseases in Portugal – a multicentre, nationwide study"

**Supplementary table 4 -** **Demographic and clinical data of patients with inflammatory and non-inflammatory RMDs with available serology.**

|  | **Overall**  **(n=79)** | **Inflammatory joint diseases^1^**  **(n=40)** | **CTD/Vasculitis^2^**  **(n=25)** | **Non-inflammatory diseases^3^**  **(n=14)** |  | **p value** |
| --- | --- | --- | --- | --- | --- | --- |
| **Age (years), median (IQR)** | 57 (14) | 53 (16) | 58 (15) | 61 (14) |  | **0.026** |
| **Female, N (%)** | 61 (77.2) | 26 (65.0) | 22 (88.0) | 13 (92.9) |  | **0.03** |
| **Treatment at COVID-19 diagnosis, N (%)^4^** | | | | | | |
| No DMARDs | 24 (30.4) | 8 (20.0) | 4 (16.0) | 12 (85.7) |  | **<0.001** |
| Glucocorticoids | 30 (38.0) | 17 (42.5) | 12 (48.0) | 1 (7.1) |  | **0.029** |
| csDMARDs | 37 (46.8) | 17 (42.5) | 18 (72.0) | 2 (14.3) |  | **0.002** |
| TNFi | 9 (11.4) | 9 (22.5) | 0 (0.0) | 0 (0.0) |  | **0.007** |
| Rituximab | 2 (2.5) | 1 (2.5) | 1 (4.0) | 0 (0.0) |  | 0.747 |
| Other b/tsDMARDs | 4 (5.1) | 3 (7.5) | 1 (4.0) | 0 (0.0) |  | 0.522 |
| **COVID-19 severity**  **(N, %)** | | | | | | 0.425 |
| Asymptomatic^5^ | 8 (10.1) | 5 (12.5) | 3 (12.0) | 0 (0.0) |  |  |
| Mild | 12 (15.2) | 8 (20.0) | 2 (8.0) | 2 (14.3) |  |  |
| Moderate | 43 (54.4) | 21 (52.5) | 15 (60.0) | 7 (50.0) |  |  |
| Severe | 15 (19.0) | 6 (15.0) | 4 (16.0) | 5 (35.7) |  |  |
| Critical | 1 (1.3) | 0 (0.0) | 1 (4.0) | 0 (0.0) |  |  |
| **Sample timing^6^**  **(days), median (IQR)** | 237 (125) | 256 (123) | 225 (161) | 234 (65) |  | 0.543 |
| **Seroconversion (N,%)** | 70 (89) | 33, 83 | 23, 92 | 14, 100 |  | 0.168 |
| **IgG titers GM±GSD (min.-max)** | 1/1508±4.075 | 1/1436±3.96  (1/100-1/25600) | 1/1367±4.262  (1/100-1/25600) | 1/1903±4.327 |  | 0.051 |

1 – Includes rheumatoid arthritis, psoriatic arthritis (PsA), spondyloarthritis other than PsA, RS3PE, undifferentiated arthritis and microcrystalline arthritis, adult onset Still disease. 2 – Includes systemic lupus erythematosus, undifferentiated connective tissue disease, mixed connective tissue disease, systemic sclerosis, Sjögren syndrome, giant cell arteritis, Behçet disease. 3 – Fibromyalgia, osteoarthritis, osteoporosis, Paget bone disease; 4 - treatments are specified when used in more than 5 patients, other drugs used include azathioprine (n=4), leflunomide (n=4), mycophenolate (n=1), sulfassalazine (n=3), belimumab (n=1), rituximab (n=2), tocilizumab (n=2), and ustekinumab (n=1). 5 – reference for statistical analysis; 6 – relative to symptom onset or first positive RT-PCR test if asymptomatic. CTD – connective tissue diseases; GM: geometric mean; GSD: geometric SD factor; TNFi: tumour necrosis factor inhibitors.
