## Supplementary table 3 for "Risk factors for infection, predictors of severe disease and antibody response to COVID-19 in patients with rheumatic diseases in Portugal – a multicentre, nationwide study"

**Supplementary table 3 – Symptoms and laboratory data of COVID-19+ patients**

| **Epidemiologic link, N (%)** | |
| --- | --- |
| Direct contact | 118 (65.9) |
| Health care | 23 (12.8) |
| Travel to high incidence countries | 4 (2.2) |
| Unknown | 34 (19.0) |
| **Symptoms, N (%)** | |
| Cough | 91 (50.8) |
| Fever | 87 (48.6) |
| Malaise | 81 (45.3) |
| Fatigue | 67 (37.4) |
| Myalgia | 61 (34.1) |
| Headache | 55 (30.7) |
| Anosmia | 55 (30.7) |
| Dysgeusia | 52 (29.1) |
| Dyspnoea | 36 (20.1) |
| Diarrhoea | 36 (20.1) |
| Thoracic pain | 32 (17.9) |
| Odynophagia | 28 (15.6) |
| Arthralgia | 27 (15.1) |
| Rhinorrhea | 25 (14) |
| Abdominal pain | 16 (8.9) |
| Vomiting | 14 (7.8) |
| **Laboratory abnormalities, n/total N** | |
| Anaemia | 6/60 |
| Leukopenia | 19/60 |
| Lymphopenia | 36/60 |
| Thrombocytopenia | 6/60 |
| Elevated transaminases | 15/59 |
| Elevated C-reactive protein | 27/59 |
| High D-dimers | 26/38 |
| Low fibrinogen | 2/30 |
| Ferritin>2000 | 4/28 |
| High IL6 | 1/7 |
