## Supplementary table 2 for "Risk factors for infection, predictors of severe disease and antibody response to COVID-19 in patients with rheumatic diseases in Portugal – a multicentre, nationwide study"

**Supplementary table 2 – Demographic and clinical characteristics of COVID-19+ patients with and without inflammatory rheumatic diseases.**

|  | **Inflammatory RMDs* (n=162)** | **Non-inflammatory RMDs* (n=17)** | **p-value** |
| --- | --- | --- | --- |
| **Female, N (%)** | 121 (74.7) | 16 (94.1) | 0.127 |
| **Age, median (IQR), in years** | 54 (19) | 64 (14) | **0.001** |
| **Caucasian, N (%)** | 136 (83.9) | 14 (82.4) | 0.697 |
| **Age at diagnosis, mean±SD, in years** | 45.3±15.7 | 60.9±10.8 | **<0.001** |
| **Diagnosis delay, median (IQR), in years** | 1.0 (4.0) | 0.5 (5.0) | 0.636 |
| **Disease duration, median (IQR), in years** | 7.5 (11.1) | 2.7 (6.1) | **0.007** |
| **Comorbidities, N (%)** |  |  |  |
| ≥ 1 comorbidity | 65 (40.1%) | 12 (70.6%) | **0.020** |
| ≥ 2 comorbidities | 27 (16.7%) | 8 (47.1%) | **0.006** |
| **Type of COVID-19 diagnosis, N (%)** |  |  | 0.608 |
| PCR confirmed | 153 (94.4) | 17 (100.0) |  |
| Positive Serology | 5 (3.1) | 0 (0.0) |  |
| Suspected | 4 (2.5) | 0 (0.0) |  |
| **COVID-19 severity, N (%)** |  |  | 0.174 |
| Asymptomatic | 17 (10.5) | 0 (0.0) |  |
| Mild | 27 (16.7) | 2 (11.8) |  |
| Moderate | 81 (50.1) | 7 (41.2) |  |
| Severe | 24 (14.8) | 6 (35.3) |  |
| Critical | 13 (8.0) | 2 (11.8) |  |
| **COVID-10 Hospitalization care, N (%)** |  |  |  |
| Hospitalization | 37 (22.8) | 8 (47.1) | **0.042** |
| Supplemental oxygen | 28 (17.3) | 6 (35.3) | 0.103 |
| Non-invasive ventilation | 11 (6.8) | 1 (5.9) | 0.910 |
| Invasive ventilation | 3 (1.9) | 1 (5.9) | 0.338 |
| **COVID-19 treatment, N (%)** |  |  |  |
| Corticosteroids | 15 (9.3) | 4 (23.5) | 0.073 |
| Hydroxychloroquine | 25 (15.4) | 3 (17.6) | 0.737 |
| Azithromycin | 13 (8.0) | 3 (17.6) | 0.189 |
| Other antibiotic | 5 (3.1) | 1 (5.9) | 0.455 |
| Lopinavir/ritonavir | 4 (2.5) | 2 (11.8) | 0.105 |
| Remdesivir | 2 (1.2) | 1 (5.9) | 0.264 |
| Tocilizumab | 1 (0.6) | 0 (0.0) | 0.743 |
| Intravenous immunoglobulin | 1 (0.6) | 0 (0.0) | 0.743 |
| **COVID-19 complications, N (%)** |  |  |  |
| Acute respiratory distress syndrome | 11 (6.8) | 2 (11.8) | 0.622 |
| Heart failure | 1 (0.6) | 0 (0.0) | 0.740 |
| Bacterial infection | 15 (9.3) | 1 (5.9) | 0.609 |
| Macrophage activation syndrome | 2 (1.2) | 0 (0.0) | 0.638 |
| Thromboembolic event | 1 (0.6) | 0 (0.0) | 0.742 |
| Acute kidney failure | 6 (3.7) | 2 (11.8) | 0.226 |

*See table 1 for diagnosis.
